# Nurture Early for Optimal Nutrition (NEON): A pilot cluster randomised controlled trial of community-facilitator-led participatory learning and action women’s groups to improve infant feeding & care among South Asian families in East London

**DOI:** 10.64898/2026.08.28.26361604

**Authors:** Logan Manikam, Asna Fatima, Priyanka Patil, Chyntia Aryanti Mayadewi, Tala El Khatib, Joanna Drazdzewska, Oyinlola Oyebode, Clare H. Llewellyn, Kelley Webb-Martin, Carol Irish, Mfon Archibong, Jenny Gilmour, Phoebe Kalungi, Neha Batura, Kalpita Shringarpure, Monica Lakhanpaul, Michelle Heys, the NEON Steering Team

## Abstract

South Asian communities in the UK experience disproportionate maternal and child health inequalities linked to non-recommended infant feeding practices, limited health literacy, and socioeconomic constraints. Participatory learning and action (PLA) is effective in low- and middle-income countries, but high-income evidence is scarce. This pilot assessed the feasibility of a community facilitator-led PLA intervention to improve infant feeding among South Asian families in East London.

A three-arm pilot feasibility cluster randomised controlled trial (ISRCTN10234623) was conducted in Tower Hamlets and Newham, East London (May–September 2022), with 12 wards randomised 1:1:1 to face-to-face PLA, online PLA, or usual care. Multilingual community facilitators delivered eight biweekly sessions over 14 weeks. Feasibility outcomes were assessed against prespecified Go/Stop criteria; exploratory outcomes included child feeding behaviours (Children’s Eating Behaviour Questionnaire, CEBQ), parental feeding style (Parental Feeding Style Questionnaire, PFSQ), and child BMI Z-scores.

Of 263 enrolled participants, 261 had a recorded trial arm allocation; consent to the pilot feasibility study was 70.7% (186/263; 95% CI 65.0–75.9%) meeting the ≥50% Go criterion. Attendance was 37% (Tower Hamlets 59%, Newham 29%), below the ≥80% Go threshold. Six-month retention was 54.8% (Tower Hamlets 78%, Newham 48.5%; 95% CI 41.8–55.3%), triggering the Definite Stop criterion. Significant baseline imbalances included BMI Z-score (p = 0.005), ethnicity, borough, and education; no between-arm BMI differences were observed at follow-up (p = 0.249). CEBQ and PFSQ baseline completion was 24.5% and 23.0%, with no usable follow-up data. PLA Phases 3 and 4 were not completed by any group; all participants providing feedback reported it acceptable.

Recruitment was feasible and the intervention acceptable, but a Definite Stop criterion was triggered in Newham, no group completed the full PLA cycle, and outcome data were insufficient for evaluation. A definitive trial requires stratified randomisation, digitised multilingual data collection, participant reimbursement, and explicit PLA phase-completion criteria.

**Trial registration:** ISRCTN10234623 (IRAS ID: 296259; Ethics Ref: 21/SW/0142).

## INTRODUCTION

The first 1,000 days of life, from conception to a child’s second birthday, represent a critical window for growth, neurodevelopment, and long-term health. Maternal and family behaviours during this period, particularly infant feeding and caregiving practices, have lasting effects on health trajectories and influence the risk of obesity, type 2 diabetes, cardiovascular disease, and dental disorders in later life [1,2]. Optimal nutrition and responsive caregiving are conversely associated with improved cognitive development, educational attainment, and psychosocial wellbeing [2].

Despite overall improvements in child health in the United Kingdom (UK), substantial ethnic health inequalities persist. South Asian (SA) communities, particularly families of Pakistani and Bangladeshi origin, experience disproportionately poorer maternal and child health outcomes compared with the wider UK population [3,4]. Evidence from qualitative and systematic investigations demonstrates continued deviation from recommended infant and young child feeding guidelines among SA families in the UK, driven by bicultural identity challenges, low acculturation, conflicting advice from healthcare professionals and family members, and socioeconomic constraints [5–8].

Participatory learning and action (PLA) is a community mobilisation approach in which trained facilitators lead structured group cycles of problem identification, strategy development, implementation, and evaluation [9,10]. This model is endorsed by the World Health Organization as an effective strategy for improving maternal and newborn health outcomes [9]. Evidence from low- and middle-income countries (LMICs) demonstrates that women’s groups practising PLA can reduce maternal and neonatal mortality, improve breastfeeding rates, and strengthen care-seeking behaviours [10–12]. PLA approaches may also hold promise in high-income countries (HICs) for addressing health inequalities among culturally and linguistically diverse populations, but rigorous evaluations in HIC settings remain scarce [13].

The Nurture Early for Optimal Nutrition (NEON) programme was developed to address this gap. Phase 1 employed community-based participatory research among British-Bangladeshi families in East London to identify drivers of and barriers to optimal infant feeding and caregiving, informing the co-development of a culturally adapted PLA intervention [14,15]. Phase 2 expanded the programme to a broader SA population and adapted delivery to include face-to-face and online formats in response to COVID-19 restrictions. The trial protocol has been published previously [16].

This pilot feasibility trial aimed to evaluate the feasibility, fidelity, and acceptability of a community facilitator-led PLA intervention for primary carers of infants aged under two years from SA communities in East London, and to inform the design of a definitive cluster randomised controlled trial (RCT).

Primary objectives were to assess recruitment, consent, attendance, retention, intervention fidelity, participant engagement, and acceptability, and to determine whether progression to a definitive trial was warranted according to prespecified Go/Stop criteria. An additional objective was to explore the feasibility and implementation characteristics of face-to-face and online delivery modalities.

Secondary objectives were to assess the completeness of outcome data collection, evaluate intervention delivery requirements, and estimate the variability of child BMI Z-scores to inform outcome selection and sample size calculations for a future definitive trial.

## Materials and Methods

### Published Protocol and Deviations

The full trial protocol has been published previously [16]. The methods described below reflect trial procedures as implemented. Four deviations from the published protocol are declared. First, the London Borough of Waltham Forest (WF) was unable to participate prior to recruitment owing to service pressures associated with the COVID-19 pandemic, reducing the study from three boroughs to two, ie., Tower Hamlets (TH) and Newham (NH) and from 18 to 12 randomised wards. Second, some sessions allocated to the face-to-face arm were delivered online when public health restrictions prevented in-person meetings; participants remained allocated to their original randomised arm regardless of delivery modality. Third, no groups completed Phases 3 (implementing solutions) or 4 (evaluating outcomes) of the PLA cycle owing to progressive declines in attendance; findings therefore relate primarily to Phases 1 and 2. Fourth, although the published protocol specified mothers and female carers as eligible participants, 14 fathers fulfilling primary caregiving roles were also recruited during implementation.

### Study Design and Setting

A three-arm pilot feasibility cluster randomised controlled trial was conducted in the London Boroughs of TH and NH between May and September 2022. The ward was the unit of randomisation. In each borough, the six wards with the highest density of SA residents were identified using 2011 Office for National Statistics Census data [17], yielding 12 study clusters. Participants were recruited into language-specific groups according to their ethnic and linguistic background. The intervention was delivered by trained multilingual community facilitators in participants’ preferred community languages.

### Randomisation and Allocation Concealment

Ward-level randomisation was undertaken by an independent member of the University College London (UCL) research team using a computer-generated random sequence (Research Randomizer) [18], stratified by borough and allocated in a 1:1:1 ratio to face-to-face PLA, online PLA, or usual care prior to participant recruitment. Face-to-face groups met in community or children’s centres; online groups were delivered via Zoom. Allocation was concealed from participants and recruiters during recruitment to minimise selection bias. Following completion of recruitment within each language group, allocation was disclosed to community facilitators (CFs) and participants by text message or email; CFs and participants were therefore necessarily unblinded to allocation. Outcome data were collected by trained community researchers (CRs) who recorded data on an electronic platform linking participant identifiers to outcome data, maintaining CR blinding to allocation where possible.

### Eligibility Criteria

Eligible participants were mothers, fathers, or other primary carers of children aged under 24 months who identified as Indian, Pakistani, Bangladeshi, or Sri Lankan, and resided within a randomised study ward in Tower Hamlets or Newham. Participants were required to provide informed consent and to communicate in English or one of the study-supported community languages (Bengali, Sylheti, Urdu, Punjabi, Gujarati, or Tamil). Individuals were excluded if they were aged under 18 years, anticipated relocating outside the study area during the intervention period, or had participated in another research study within the preceding four weeks.

### Sample Size

As a pilot feasibility trial, the study was not powered to detect differences in effectiveness outcomes. The sample size was determined to estimate key feasibility parameters with sufficient precision to inform a future definitive trial. The published protocol proposed 20–32 PLA groups of 6–8 participants, yielding an anticipated 288–384 participants. Assuming 80% retention, this was estimated to provide 85% confidence that the true retention rate would fall within ±4 percentage points of the observed rate, based on a geometric mean cluster size of 16, a coefficient of variation of 0.70, and an Intraclass Correlation Coefficient (ICC) of 0.02 [16]. In practice, 24 intervention PLA groups were delivered (10 face-to-face, 14 online); usual care participants received no PLA sessions.

### Recruitment

Participants were recruited using three complementary strategies: community facilitator (CF)-led snowball recruitment through community networks; referrals from health visitors, general practitioners, and midwives serving the study wards; and social media advertising via Facebook and Instagram. All study materials were available in English and the study-supported community languages. Recruiters remained unaware of ward allocation during recruitment to minimise selection bias (Fig 1).

**Fig 1.**
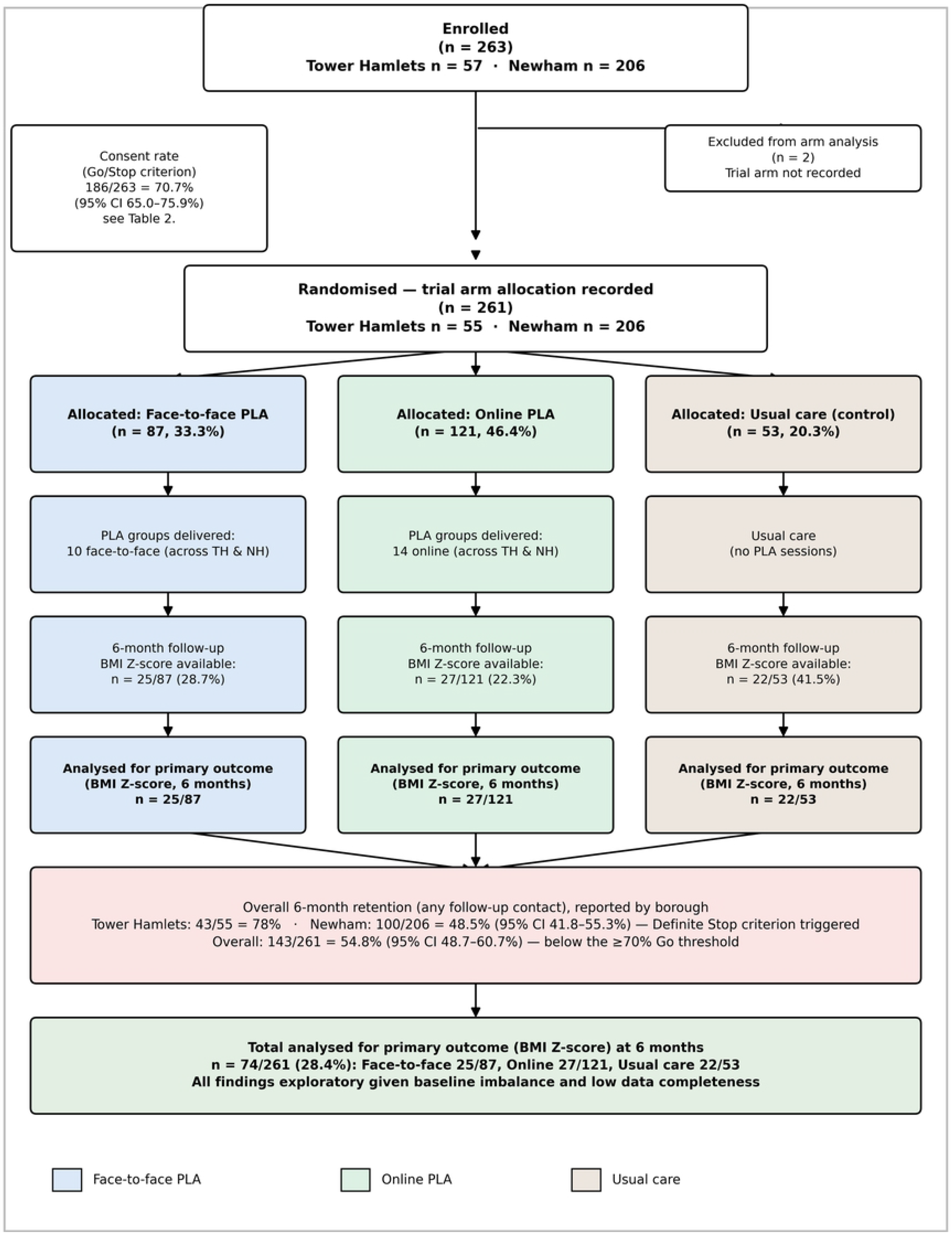
CONSORT flow diagram for the NEON pilot feasibility trial. Enrolment, randomisation, allocation, and follow-up are shown for each of the three trial arms (face-to-face PLA, online PLA, usual care), including consent rate, six-month retention by borough, and the number analysed for the primary outcome (BMI Z-score).

### Patient and Public Involvement

Patient and public involvement was embedded throughout. Five community researchers (CRs) from Phase 1 of the NEON programme, representing diverse SA communities in East London, contributed to study design, protocol development, participant recruitment, data collection, interpretation of findings, and dissemination. Ten additional CFs were recruited for Phase 2. All CRs and CFs received structured pre-intervention training delivered by Women and Children First, covering PLA methodology, group facilitation, safeguarding, and cultural competence.

### Intervention: NEON Women’s Group PLA Cycle

Each PLA cycle comprised eight fortnightly sessions over 14 weeks, following four structured phases: (1) identifying and prioritising infant feeding and care issues; (2) identifying barriers and designing solutions; (3) implementing solutions; and (4) evaluating outcomes. All sessions followed standardised topic guides. A culturally adapted toolkit, co-developed with South Asian community facilitators and independent health experts during Phase 1, comprised: a facilitator manual; picture cards illustrating recommended and non-recommended feeding practices; healthy baby food recipes; community asset maps; and local resource lists [16].

### Control Arm: Usual Care

Control arm participants received standard National Health Service (NHS) maternal and child health services from health visitors and general practitioners, including scheduled postnatal and early childhood visits at birth, 6–8 weeks, 12–16 weeks, one year, and 2–2.5 years, covering child growth, development, immunisation, and infant feeding guidance, without any PLA activities.

### Data Collection

Primary outcome and questionnaire data were collected at three time points: baseline (pre-intervention), 14 weeks (end of PLA cycle), and six-month follow-up. The planned 12-month follow-up was removed owing to COVID-19 restrictions. Process evaluation data (attendance, fidelity, and participant feedback) were collected at the end of each fortnightly session throughout the intervention period. Child weight (kg) and height or recumbent length (cm) were measured by CRs using calibrated digital scales and length boards; BMI Z-scores were calculated using WHO 2006 child growth standards adjusted for age and sex [19]. The CEBQ and PFSQ were administered by CRs with facilitator-assisted verbal translation for participants with limited English literacy. Administration was paper-based and contingent on session attendance; this constituted a deviation from the protocol specification of digitised multilingual administration independent of PLA sessions.

### Outcome Measures

Feasibility outcomes including consent rate, session attendance, six-month retention, intervention support (participant ratings of session content, frequency, duration, and quality), and acceptability were assessed against prespecified Go/Stop progression criteria [16]. Child feeding behaviours were measured using the CEBQ [20,21], adapted for use with infants, comprising eight domains rated on a five-point Likert scale (1 = Never to 5 = Always): Food Responsiveness, Enjoyment of Food, Emotional Overeating, Desire to Drink, Satiety Responsiveness, Slowness in Eating, Emotional Undereating, and Food Fussiness. The published protocol planned collection of six of eight CEBQ domains; all eight were administered in practice, constituting a minor protocol deviation. Parental feeding style was assessed using the PFSQ as originally developed by Daniels et al [22] and as described in the published protocol [16], measuring four domains on a five-point Likert scale: Emotional Feeding, Instrumental Feeding, Encouragement to Eat, and Control over Eating.

Mealtime video recordings were collected from intervention arm participants and coded using ELAN Linguistic Annotator software for six behaviour themes derived from the NEON Phase 1 formative study [14,23]: early and late introduction of semi-solid/solid foods, forced feeding, distraction feeding, preferences for milk and sweet foods, chasing fussy eaters, and prolonged hand and spoon feeding.

Sustainability capacity was assessed using a nine-domain structured framework described in the published protocol [16] (group leadership, structure, problem assessment, resource mobilisation, links to other groups, critical awareness, relationship with outside agents, programme management, and participant engagement), scored from structured reflection forms completed by CFs and CRs after each PLA cycle (Fig 2). An Equality Impact Assessment (EIA) evaluated the likely effects of the intervention across protected characteristics as defined by the UK Equality Act 2010. Implementation fidelity was assessed by CRs acting as independent observers using a structured checklist evaluating session structure, planned activities, participant engagement, and PLA phase completion on a four-point scale (1 = minimal adherence; 4 = full adherence).

**Fig 2.**
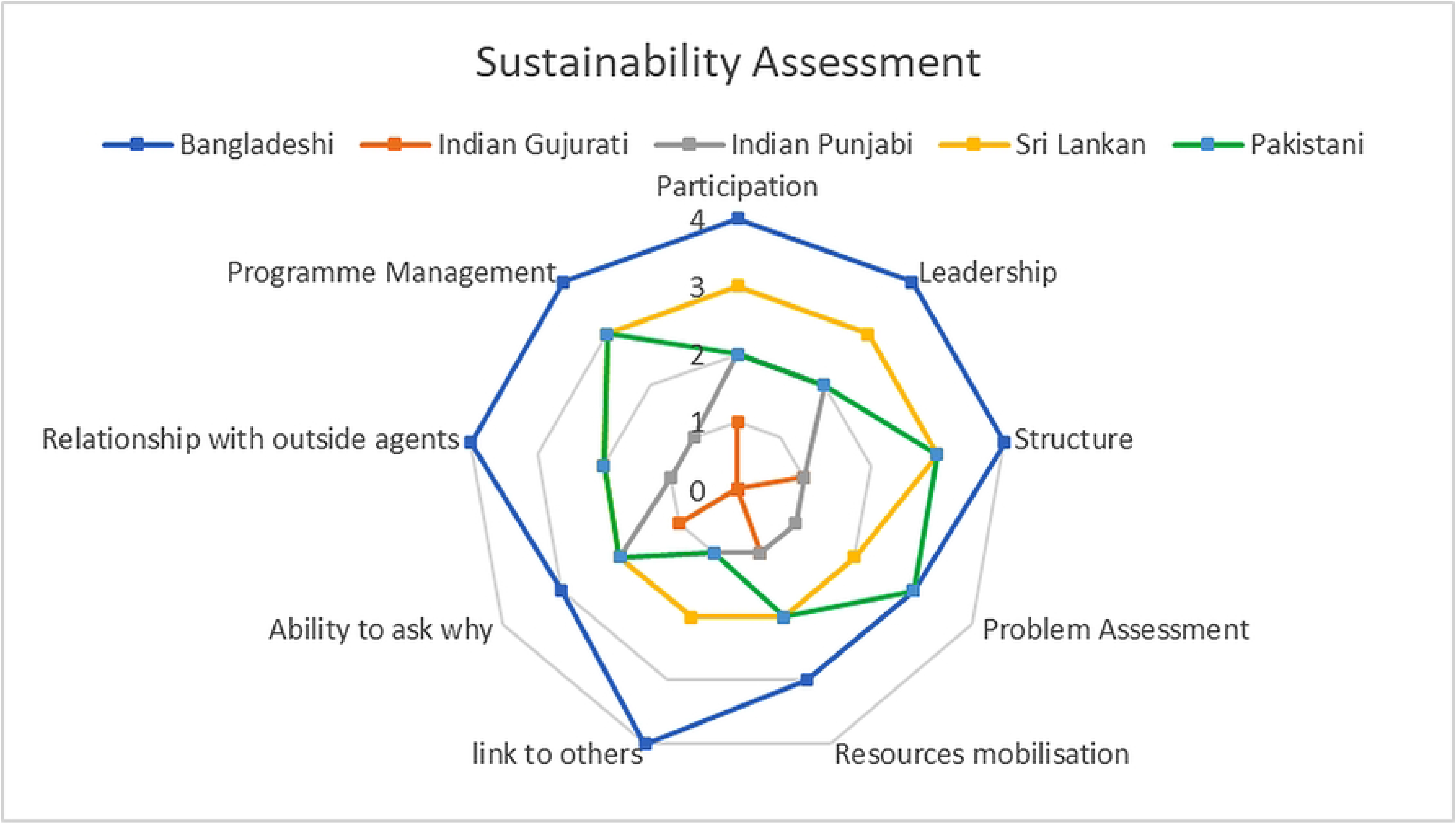
Sustainability assessment scores by ethnic group across nine domains. Scores range 0–4 (0 = minimal adherence, 4 = full adherence), covering intervention structure, planned activities, participant engagement, and PLA phase completion.

### Statistical Analysis

Consistent with the published protocol [16], this pilot trial was designed to assess feasibility rather than to test effectiveness hypotheses; no formal power calculation for effectiveness outcomes was performed and no inferential efficacy testing was pre-specified. Feasibility outcomes (consent rate, attendance, and retention) were summarised as proportions with 95% confidence intervals (CIs) calculated using the Wilson score method. Continuous outcomes (BMI Z-scores, CEBQ and PFSQ domain scores) were summarised using means and standard deviations by trial arm and time point; questionnaire analyses are restricted to baseline data given the absence of usable follow-up responses. Between-arm differences in BMI Z-scores at each time point were examined using one-way analysis of variance (ANOVA); within-arm changes from baseline to six-month follow-up were examined using the Wilcoxon signed-rank test, restricted to participants with complete paired data. The ICC for BMI Z-score was estimated from intervention arm PLA groups using the one-way ANOVA method. All analyses were descriptive; results were assessed collectively against the prespecified Go/Stop criteria [16].

### Ethics

Ethics approval was obtained from the UCL Research Ethics Committee and the NHS Health Research Authority (IRAS ID: 296259). Informed consent was obtained from all participants prior to enrolment, provided in writing or by audio recording at least 24 hours after receipt of the participant information sheet in English or the participant’s preferred community language. Participation was voluntary; participants were free to withdraw at any time without prejudice to their usual care. The trial is registered with the International Standard Randomised Controlled Trial registry (ISRCTN10234623).

## Results

Results are reported in line with the published protocol[16] and assessed against prespecified Go/Stop progression criteria.

### Participant Characteristics

Of 263 enrolled participants, 261 had a recorded trial arm allocation (Fig 1): 87 (33.3%) face-to-face, 121 (46.4%) online, and 53 (20.3%) usual care. Carers comprised 246 mothers (94.3%), 14 fathers (5.4%), and one grandmother (0.4%); mean carer age was 32.0 years (SD 5.98). The majority resided in Newham (NH; 78.9%); the remainder in Tower Hamlets (TH; 21.1%). Of these, 57.9% were Bangladeshi, 69.7% first-generation migrants, 67.4% were engaged in home duties and childcare, and 24.1% had little or no English literacy. Significant baseline imbalances were identified in borough (p = 0.008), ethnicity (p = 0.002), education (p = 0.044), and BMI Z-score (p = 0.005); full characteristics are presented in Table 1.

**Table 1.** Baseline characteristics of enrolled participants by trial arm.

| Characteristic | Category | Face-to-face (n = 87) | Online (n = 121) | Usual care (n = 53) | Total (n = 261) | p-value |
| --- | --- | --- | --- | --- | --- | --- |
| **Participant demographics** |  |  |  |  |  |  |
| **Enrolled** | n | 87 | 121 | 53 | 261 | — |
| **Carer age (years)** | Mean (SD) | 32.5 (5.8) | 31.8 (6.0) | 32.0 (6.2) | 32.0<br>(5.98) | — |
| **Relationship to infant** | Mother, n (%) | 82<br>(94.3%) | 112<br>(92.6%) | 52<br>(98.1%) | 246<br>(94.3%) | — |
|  | Father, n (%) | 4 (4.6%) | 9 (7.4%) | 1 (1.9%) | 14 (5.4%) |  |
|  | Grandmother, n (%) | 1 (1.1%) | 0 (0.0%) | 0 (0.0%) | 1 (0.4%) |  |
| **Marital status** | Married, n (%) | 82<br>(94.3%) | 119<br>(98.3%) | 49<br>(92.5%) | 250<br>(95.8%) | — |
|  | Not married, n (%) | 5 (5.7%) | 2 (1.7%) | 4 (7.5%) | 11 (4.2%) |  |
| **Generation** | First generation | 60<br>(69.0%) | 88<br>(72.7%) | 34<br>(64.2%) | 182<br>(69.7%) | — |
|  | Second generation | 18<br>(20.7%) | 22<br>(18.2%) | 16<br>(30.2%) | 56<br>(21.5%) |  |
|  | Third+ generation | 2 (2.3%) | 1 (0.8%) | 0 (0.0%) | 3 (1.1%) |  |
|  | Not recorded | 7 (8.0%) | 10 (8.3%) | 3 (5.7%) | 20 (7.7%) |  |
| ***Study setting*** |  |  |  |  |  |  |
| **Borough** | Newham, n (%) | 59<br>(67.8%) | 103<br>(85.1%) | 44<br>(83.0%) | 206<br>(78.9%) | 0.008 |
|  | Tower Hamlets, n (%) | 28<br>(32.2%) | 18<br>(14.9%) | 9 (17.0%) | 55<br>(21.1%) | 0.008 |
| ***Ethnicity*** |  |  |  |  |  |  |
| **Ethnicity** | Bangladeshi | 62<br>(71.3%) | 59<br>(48.8%) | 30<br>(56.6%) | 151<br>(57.9%) | 0.002 |
|  | Pakistani | 8 (9.2%) | 24<br>(19.8%) | 9 (17.0%) | 41<br>(15.7%) |  |
|  | Indian Gujarati | 9 (10.3%) | 11 (9.1%) | 11<br>(20.8%) | 31<br>(11.9%) |  |
|  | Indian Punjabi | 7 (8.0%) | 8 (6.6%) | 2 (3.8%) | 17 (6.5%) |  |
|  | Sri Lankan | 0 (0.0%) | 11 (9.1%) | 1 (1.9%) | 12 (4.6%) |  |
|  | Indian Tamil | 0 (0.0%) | 6 (5.0%) | 0 (0.0%) | 6 (2.3%) |  |
|  | Indian Bengali | 1 (1.1%) | 2 (1.7%) | 0 (0.0%) | 3 (1.1%) |  |
| **Infant characteristics** |  |  |  |  |  |  |
| **Infant age (months)** | Mean (SD), n = 155** | — | — | — | 12.4<br>(5.19) | — |
| **Infant sex (n = 132)\*** | Male, n (%) | 16<br>(42.1%) | 21<br>(36.2%) | 20<br>(55.6%) | 57<br>(43.2%) | — |
|  | Female, n (%) | 22<br>(57.9%) | 37<br>(63.8%) | 16<br>(44.4%) | 75<br>(56.8%) |  |
| **Socioeconomic characteristics** |  |  |  |  |  |  |
| **Education** | No qualifications | 20<br>(23.0%) | 20<br>(16.5%) | 7 (13.2%) | 47<br>(18.0%) | 0.044 |
|  | GCSE level | 33<br>(37.9%) | 52<br>(43.0%) | 20<br>(37.7%) | 105<br>(40.2%) |  |
|  | A-level | 4 (4.6%) | 21<br>(17.4%) | 6 (11.3%) | 31<br>(11.9%) |  |
|  | Higher degree | 30<br>(34.5%) | 28<br>(23.1%) | 20<br>(37.7%) | 78<br>(29.9%) |  |
| **Employment** | Home/childcare | 62<br>(71.3%) | 78<br>(64.5%) | 36<br>(67.9%) | 176<br>(67.4%) | — |
|  | Full-time | 18<br>(20.7%) | 24<br>(19.8%) | 8 (15.1%) | 50<br>(19.2%) |  |
|  | Part-time | 4 (4.6%) | 14<br>(11.6%) | 7 (13.2%) | 25 (9.6%) |  |
|  | Student | 0 (0.0%) | 2 (1.7%) | 0 (0.0%) | 2 (0.8%) |  |
|  | Not recorded | 3 (3.4%) | 3 (2.5%) | 2 (3.8%) | 8 (3.1%) |  |
| **English literacy** | Fluent | 36<br>(41.4%) | 54<br>(44.6%) | 30<br>(56.6%) | 120<br>(46.0%) | — |
|  | Moderate | 28<br>(32.2%) | 36<br>(29.8%) | 14<br>(26.4%) | 78<br>(29.9%) |  |
|  | Little to none | 23<br>(26.4%) | 31<br>(25.6%) | 9 (17.0%) | 63<br>(24.1%) |  |
| **Primary outcome at baseline** |  |  |  |  |  |  |
| **BMI Z-score** | n with data (%) | 30<br>(34.5%) | 45<br>(37.2%) | 26<br>(49.1%) | 101<br>(38.7%) | 0.005 |
|  | Mean (SD) | 0.87<br>(1.32) | −0.40<br>(1.98) | −0.11<br>(1.21) | 0.05<br>(1.70) |  |

Note: GCSE = General Certificate of Secondary Education; A-level = Advanced Level (UK pre-university qualification). SD = standard deviation. Carer age SD values do not carry percentage signs. Statistically significant baseline imbalances were identified for borough ( $p = 0.008$ ), ethnicity ( $p = 0.002$ ), education ( $p = 0.044$ ), and BMI Z-score ( $p = 0.005$ ); these limit between-arm comparisons of outcomes. p-values derived from chi-squared tests (categorical variables) and one-way ANOVA (BMI Z-score). Cells show n (%) unless otherwise stated.
\* Infant sex recorded for 132 of 261 participants; percentages are of those with recorded sex within each arm.
\*\* Infant age recorded for 155 of 261 participants; lower n reflects incomplete date-of-birth recording rather than loss to follow-up.

### Feasibility Outcomes

The consent rate of 70.7% (186/263; 95% CI 65.0–75.9%) met the ≥50% Go criterion. Session attendance was 37% (95% CI 30.5–44.2%), below the ≥80% Go threshold (TH 59%, NH 29%). Six-month retention was 54.8% (143/261; 95% CI 48.7–60.7%), calculated as the sum of borough-level retained participants; TH retention was 78% (43/55; 95% CI 65.6–87.1%), and NH retention was 48.5% (100/206; 95% CI 41.8–55.3%), triggering the Definite Stop criterion. TH groups demonstrated high fidelity with all planned sessions delivered (some online owing to COVID-19 restrictions); NH groups showed variable fidelity. PLA Phases 3 and 4 were not completed by any group. All 46 participants completing feedback questionnaires (24.7% of the intervention arm) reported the intervention as acceptable; session length and limited English literacy were identified as recurring barriers (Table 2).

**Table 2.** Prespecified Go/Stop criteria and observed results for the NEON pilot feasibility RCT.

| Feasibility criterion | Definite Go | Definite Stop | Observed result | Assessment |
| --- | --- | --- | --- | --- |
| **Consent rate** | $\geq 50\%$ of eligible participants consenting to pilot feasibility study | $< 40\%$ of eligible participants consenting to pilot feasibility trial | **70.7% (186/263; 95% CI 65.0–75.9%)**<br>186 of 263 enrolled participants consented (263 enrolled is used here as the denominator for eligible participants approached, as a separate pre-enrolment eligibility screening log was not maintained); 261 of these had a recorded arm allocation (Table 1). | **✓ Go criterion met** |
| **Session attendance** | $\geq 80\%$ of mothers<sup>1</sup> attend $\geq 60\%$ of planned sessions in the intervention arm | $< 20\%$ of mothers<sup>1</sup> attend $\geq 60\%$ of sessions as planned in each intervention arm | **37% overall (95% CI 30.5–44.2%)**<br>Tower Hamlets: 59% \| Newham: 29%<br>Note: reported as mean session attendance rate per session; individual-level $\geq 60\%$ session-completion data were not available. | **Below Go; above Stop** |
| **Six-month retention** | Retention of $\geq 70\%$ of consented | Retention of $< 50\%$ of | **54.8% overall (143/261; 95% CI 48.7–60.7%)<sup>†</sup>** | **Below Go overall** |
|  | participants for primary outcome data collection | participants at 6 months | Tower Hamlets: 78% (43/55) \| Newham: 48.5% (100/206; 95% CI 41.8–55.3%)<sup>3</sup> |  |
| **Intervention support** | High intervention support with respect to content, frequency, duration and quality | Low support of intervention procedures | Tower Hamlets: high fidelity; all 8 planned sessions delivered (some online per COVID-19 restrictions).<br>Newham: variable fidelity. PLA Phases 3 and 4 not completed by any group.<br>24 PLA groups established (10 face-to-face; 14 online). | **Below Go; above Stop** |
| **Acceptability** | Intervention is perceived as acceptable | Intervention perceived as unacceptable | 46/186 intervention participants (24.7%) completed feedback questionnaires.<br>All 46 reported the intervention as acceptable.<br>Recurring barriers: session length; limited English literacy. | **✓ Go criterion met** |

<sup>†</sup> Overall six-month retention is calculated as the sum of the two borough-level retained counts
(Tower Hamlets 43/55 + Newham 100/206 = 143/261 = 54.8%; 95% CI 48.7–60.7%, Wilson score method)
<sup>1</sup> "Mothers" includes eligible female carers, consistent with trial eligibility criteria (mothers or female carers of an infant aged <24 months).

### Session Attendance and Groups

Twenty-four intervention PLA groups were established (10 face-to-face, 14 online). Each planned eight fortnightly sessions over 14 weeks; mean attendance was 37% overall (TH: 59%; NH: 29%). Facilitators prioritised core PLA content in response to variable attendance, as reported in facilitator feedback forms.

### Acceptability and Participant Feedback

Feedback questionnaires were completed by 46 of 186 intervention participants (24.7%). All 46 reported the intervention as acceptable. Six themes emerged: (1) value of culturally tailored content and community language delivery; (2) social support and peer relationships; (3) trust in community facilitators; (4) session length experienced as lengthy; (5) interest in translated written materials; and (6) connectivity difficulties among online participants.

#### Implementation Sustainability Assessment

Sustainability capacity was assessed across nine domains scored 0–4 (Fig 2). The Bangladeshi group scored highest overall (mean 2.6/4), achieving the only maximum score (4/4) on Participation. The Sri Lankan group ranked second (mean 1.7/4). Programme Management was consistently the highest-scoring domain across groups; the four problem-solving domains (Problem Assessment, Resource Mobilisation, Links to Other Groups, Critical Awareness) were consistently lowest. The Indian Punjabi and Indian Gujarati groups recorded uniformly low scores (means 1.1 and 0.7/4 respectively).

#### Exploratory Outcome Measures

##### Child Feeding Behaviour (CEBQ)

The CEBQ was completed at baseline by 64 participants (24.5%; 95% CI 19.7–30.1%); a small number of follow-up questionnaires were returned but all were blank, so no usable follow-up CEBQ data were obtained, precluding longitudinal analysis. Between-arm comparisons were not conducted. Domain scores ranged from 2.46 (Emotional Overeating; 95% CI 2.22–2.70) to 3.21 (Satiety Responsiveness; 95% CI 3.01–3.41); Food Responsiveness (2.65) and Desire to Drink (2.71) were below the scale midpoint. The Slowness in Eating domain used a two-item adaptation (Table 3).

**Table 3.** Children’s Eating Behaviour Questionnaire (CEBQ) baseline domain means.

| CEBQ Domain | Construct | n | Mean | SD | 95% CI |
| --- | --- | --- | --- | --- | --- |
| **Satiety Responsiveness (SR)** | Sensitivity to fullness | 59 | 3.21 | 0.77 | 3.01–3.41 |
| **Slowness in Eating (SE) \*** | Eating pace | 61 | 3.16 | 0.96 | 2.92–3.40 |
| **Enjoyment of Food (EF)** | Positive food engagement | 61 | 3.15 | 0.89 | 2.93–3.37 |
| **Emotional Undereating (EUE)** | Avoidance when distressed | 61 | 3.13 | 0.91 | 2.90–3.36 |
| **Food Fussiness (FF)** | Avoidance of novel foods | 61 | 2.86 | 0.65 | 2.70–3.02 |
| **Desire to Drink (DD)** | Drink-seeking behaviour | 59 | 2.71 | 0.91 | 2.48–2.94 |
| **Food Responsiveness (FR)** | Appetite-driven eating | 62 | 2.65 | 0.68 | 2.48–2.82 |
| **Emotional Overeating (EOE)** | Food as emotional response | 61 | 2.46 | 0.97 | 2.22–2.70 |

Note: CEBQ items scored on a 5-point Likert scale (1 = Never to 5 = Always). Higher scores indicate greater frequency of the behaviour. 95% CIs calculated as mean ± 1.96 × SE.

##### Parental Feeding Style (PFSQ)

The PFSQ was completed at baseline by 60 participants (23.0%; 95% CI 18.3–28.5%); only one partial follow-up response was recorded (7 of 26 items), insufficient to support any follow-up analysis. Encouragement to Eat was the highest-scoring domain (mean 3.93, 95% CI 3.75–4.11); Instrumental Feeding (3.07) and Emotional Feeding (2.93) were both below the scale midpoint. The Control over Eating domain was scored without validated reverse coding (Table 4).

**Table 4.** Parental Feeding Style Questionnaire (PFSQ) baseline domain means.

| PFSQ Domain | Construct | n | Mean | SD | 95% CI |
| --- | --- | --- | --- | --- | --- |
| **Encouragement to Eat (EE)** | Positive promotion of eating | 59 | 3.93 | 0.72 | 3.75–4.11 |
| **Control over Eating (CE)** | Parental control of feeding | 59 | 3.34 | 0.66 | 3.17–3.51 |
| **Instrumental Feeding (InF)** | Food as reward or punishment | 58 | 3.07 | 1.08 | 2.79–3.35 |
| **Emotional Feeding (EmF)** | Food used for comfort/distress | 58 | 2.93 | 0.97 | 2.68–3.18 |

Note: PFSQ items scored on a 5-point Likert scale (1 = Never to 5 = Always). Higher scores indicate greater frequency of the behaviour. 95% CIs calculated as mean ± 1.96 × SE

#### Child BMI Z-Scores

BMI Z-score data were available for 101 of 261 participants (38.7%) at baseline, 101 (38.7%) at mid-intervention, and 74 (28.4%) at six-month follow-up. A statistically significant between-arm difference was identified at baseline (F(2,98) = 5.58, p = 0.005), with the face-to-face arm having a higher mean BMI Z-score (0.87, SD 1.32) than online (−0.40, SD 1.98) and usual care (−0.11, SD 1.21). Between-arm differences persisted at mid-intervention (F(2,98) = 6.80, p = 0.002) but not at six-month follow-up (F(2,71) = 1.42, p = 0.249). Within-arm Wilcoxon signed-rank tests showed no significant change in any arm (all p ≥ 0.117): face-to-face −0.49 (W = 81, p = 0.230); online +0.29 (W = 49.5, p = 0.117); usual care +0.25 (W = 56, p = 0.535). All findings are exploratory given the significant baseline imbalance and low data completeness (Table 5).

**Table 5.**
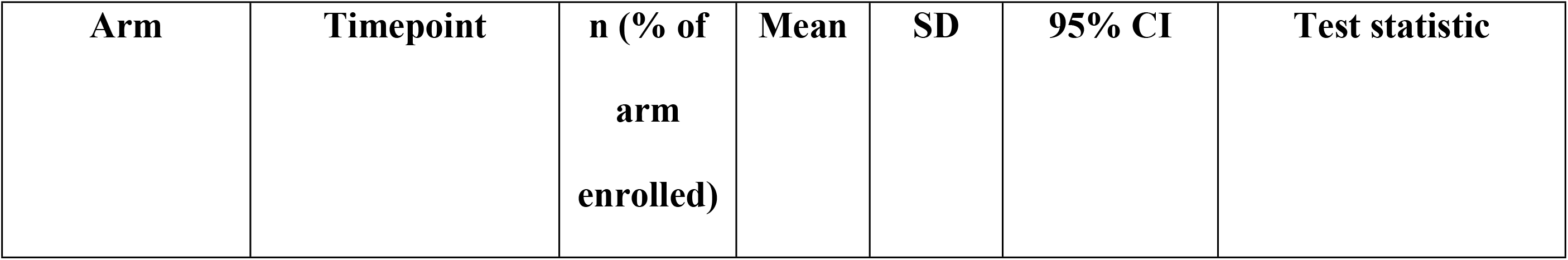
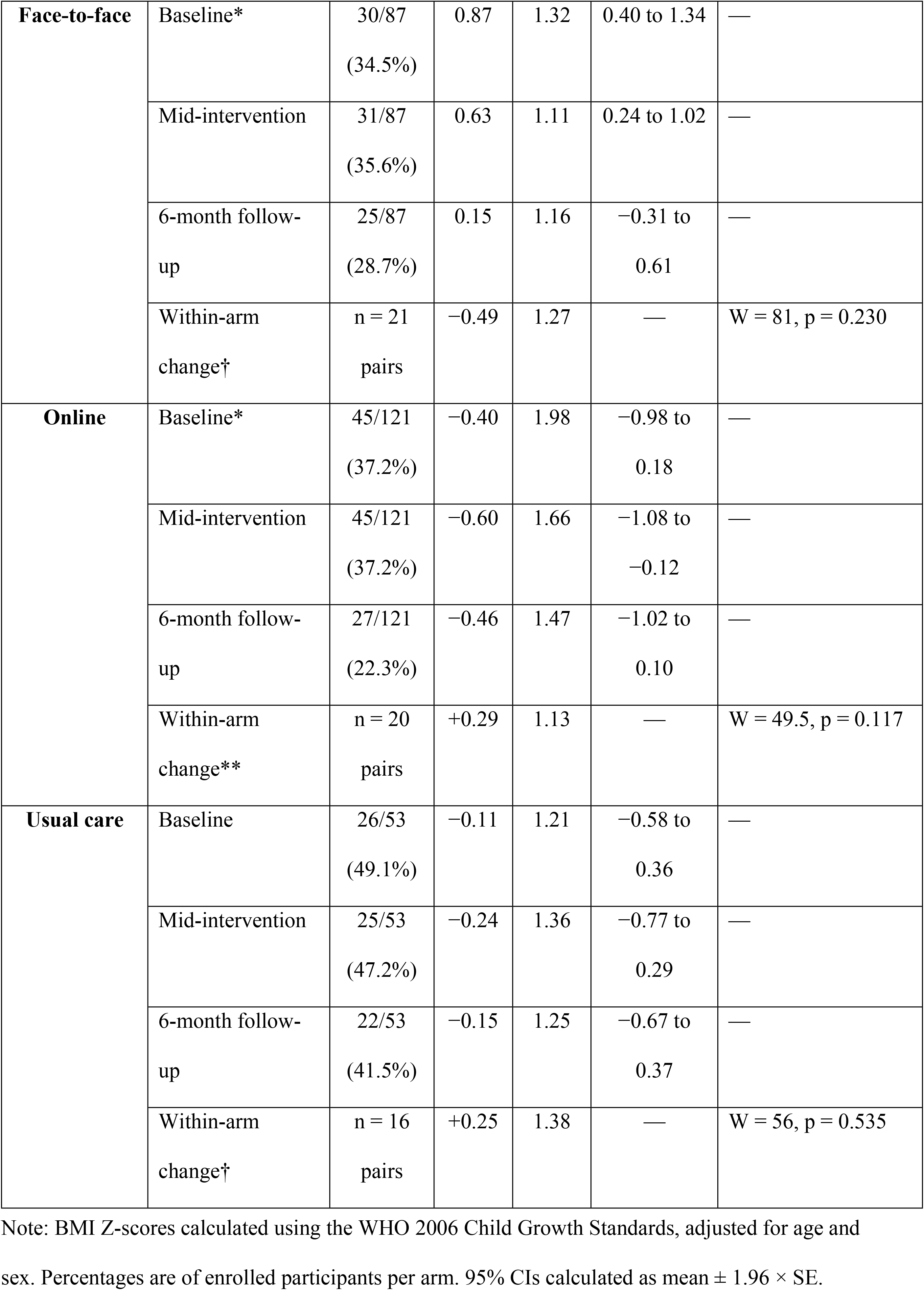
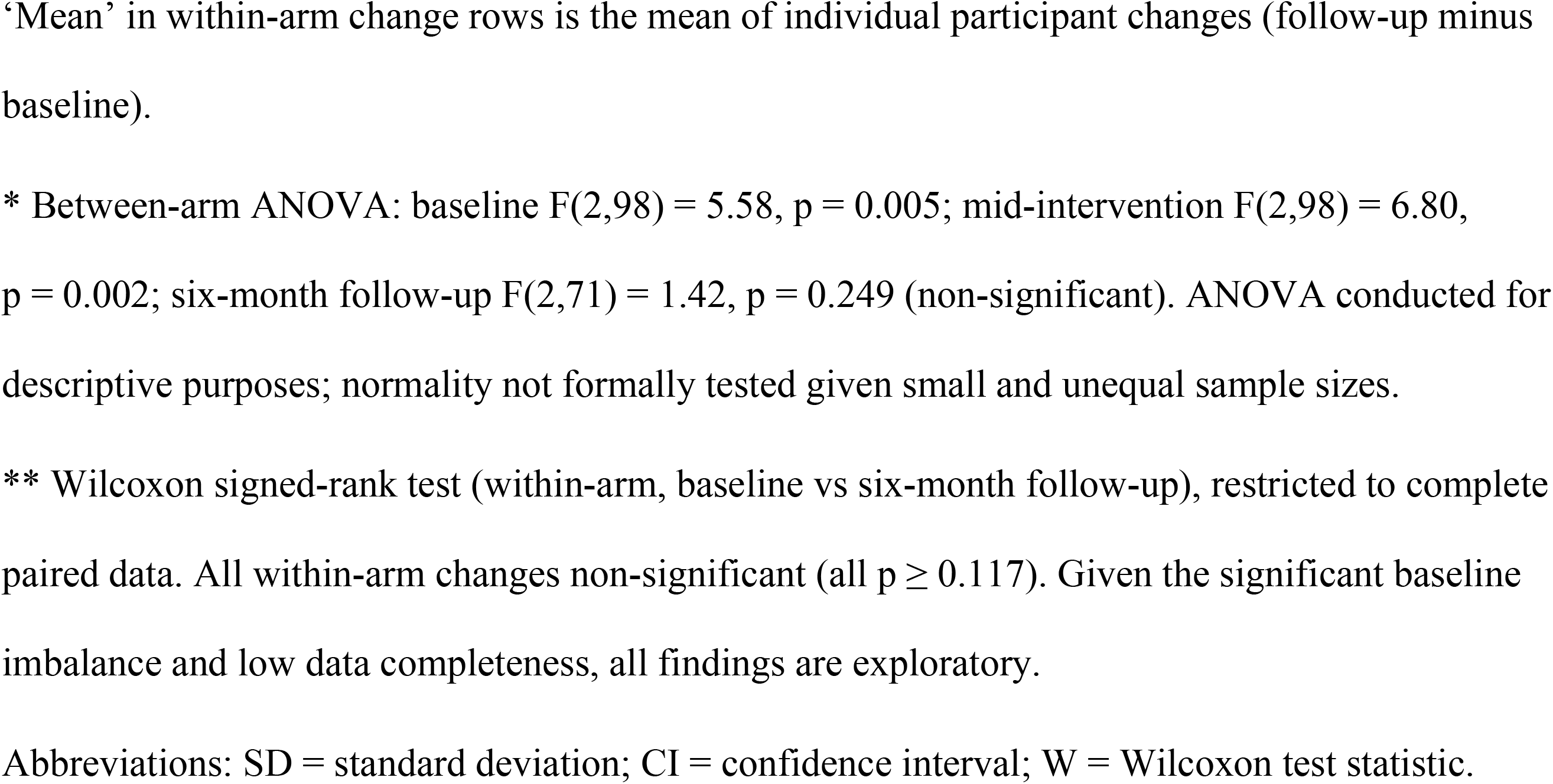
Child BMI Z-scores by trial arm and timepoint.

#### Video Observations of Feeding Behaviours

Twenty-seven mealtime videos were coded using ELAN software **[14]** (15 pre-intervention, 12 at approximately 14 weeks) across six behaviour themes (Fig 3). Forced feeding was observed in six videos (five pre, one post), exclusively among online participants; it was absent from all face-to-face sessions. Proportional changes were most notable in forced feeding (33% to 8%) and prolonged hand and spoon feeding (40% to 25%); distraction feeding was stable (47% pre, 50% post). Unequal denominators and absence of inter-rater reliability preclude inferential comparison; findings are exploratory and descriptive only.

**Fig 3.**
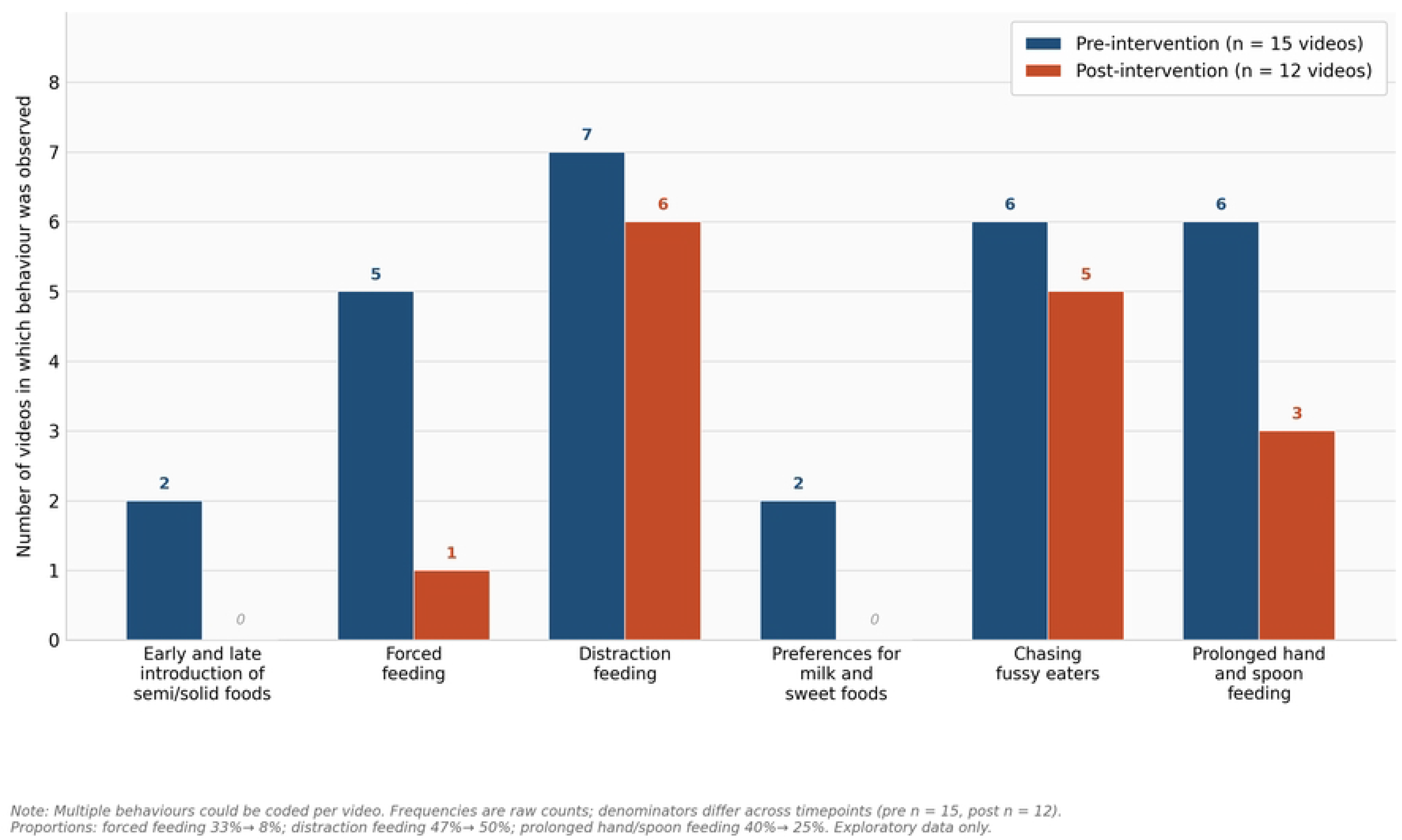
Frequency of observed feeding behaviour themes before and after the NEON PLA intervention. Twenty-seven mealtime videos were coded (15 pre-intervention, 12 post-intervention, approximately 14 weeks later); multiple behaviours could be coded per video and denominators differ across timepoints (pre n = 15, post n = 12). Proportional changes: forced feeding 33% to 8%; distraction feeding 47% to 50%; prolonged hand/spoon feeding 40% to 25%. Findings are exploratory and descriptive only.

### Additional Secondary Outcomes

Network diffusion via the eRedbook platform could not be evaluated owing to connectivity failures and low engagement. The four-day food diary was returned by fewer than ten participants, impeded by the literacy demands of prospective dietary recording given that 24.1% of participants had little or no English literacy (Table 1). Both outcomes require substantial adaptation in any definitive trial.

### Equality Impact Assessment (EIA)

The EIA identified no areas of adverse impact. Two recommendations were identified for a definitive trial: inclusion of all primary carers regardless of sex (14 fathers were enrolled despite female-only eligibility; Table 1), and proactive accessibility planning for participants with disabilities.

### Sample Size Estimation for a Definitive Trial

The intracluster correlation coefficient (ICC) for BMI Z-score at follow-up was estimated at 0.024 (95% CI −0.28 to 0.38) based on 24 intervention PLA groups, although the confidence interval was wide, indicating considerable uncertainty. The weighted pooled SD was 1.31. Using the standard two-sample formula (80% power, α = 0.05) to detect meaningful difference 0.5 of BMI Z-score units [24], we obtained an unadjusted estimate of 108. After applying inflation factor, the participant per arm increases to 127 (DEFF 1.18) and 159 per arm after 20% attrition allowance (318 in total). These estimates are preliminary given the low data completeness, baseline imbalance, and wide ICC confidence interval; a larger pilot phase is recommended to refine these parameters before a definitive trial.

### Harms

No serious adverse events were reported during the trial.

## Discussion

This pilot demonstrates that a community PLA intervention is deliverable within ethnically diverse SA communities in a HIC setting, with community-embedded multilingual recruitment meeting the prespecified consent threshold. However, the findings collectively indicate that progression to a definitive trial is not currently warranted: the Definite Stop criterion was triggered in Newham, no group completed the full PLA cycle, significant baseline imbalances preclude effectiveness inference, and outcome data completeness was insufficient across all exploratory measures. The primary contribution of this pilot is a structured characterisation of what must change before a definitive trial.

The consent rate of 70.7% (95% CI 65.0–75.9%) exceeded the ≥50% Go criterion, confirming that multilingual recruitment through facilitator networks, health visitor referral, and social media is achievable in this population. Overall session attendance of 37%, however, fell below the ≥80% Go threshold. The marked contrast between TH (59%) and NH (29%) reflects borough-level structural differences: TH benefited from established facilitator networks and geographic concentration, while NH faced greater ethnic and linguistic diversity, appointment-based referral pathways, and a higher proportion of online delivery. TH attendance was at the lower end of face-to-face attendance rates reported in comparable participatory nutrition programmes in high-income countries (50–70%) [13], NH attendance fell below these ranges. The participant profile compounds engagement challenges: 69.7% were first-generation migrants, 67.4% were engaged in home duties and childcare, and 24.1% had little or no English literacy. Six-month retention of 48.5% in NH (100/206; 95% CI 41.8–55.3%) triggered the Definite Stop criterion; TH retention of 78% met the Go criterion. Participants received no financial reimbursement; provision of reimbursement and formally validated translated instruments are essential conditions for a definitive trial.

The statistically significant baseline imbalances in BMI Z-score (F(2,98) = 5.58, p = 0.005), ethnicity (χ² = 31.55, p = 0.002), borough (χ² = 9.78, p = 0.008), and education (χ² = 12.95, p = 0.044) represent a critical methodological finding. Ward-level cluster randomisation with a small number of clusters cannot guarantee individual-level covariate balance [25]. The absence of significant between-arm differences at six-month follow-up (F(2,71) = 1.42, p = 0.249) and non-significant within-arm changes (all p ≥ 0.117) cannot therefore be interpreted as evidence for or against intervention effectiveness. A definitive trial must employ stratified randomisation by borough, ethnicity, and baseline BMI Z-score, with pre-specified covariate-adjusted primary analyses.

The non-completion of PLA Phases 3 (implementing solutions) and 4 (evaluating outcomes) by any group represents a critical fidelity gap: no participant received a complete PLA cycle as designed, and the active behaviour-change component of the intervention was never reached. This is consistent with the attendance pattern; at 37% of planned sessions, cumulative attendance was insufficient to sustain iterative PLA progression. Future trials must establish explicit phase-completion criteria and consider whether eight fortnightly sessions over 14 weeks are sufficient for communities with competing caregiving demands.

CEBQ and PFSQ baseline completion rates of 24.5% and 23.0% respectively, and the complete absence of usable follow-up data, represent a systemic data collection failure attributable to paper-based administration contingent on session attendance, which was incompatible with the literacy profile of the sample and the online delivery format. Two instrument deviations are noted: the Slowness in Eating domain used a two-item adaptation, and the Control over Eating domain was scored without the validated reverse coding; both are declared in the Results and must be addressed in future data collection. Notwithstanding these limitations, the baseline profiles provide useful context: the high Encouragement to Eat score (mean 3.93) is consistent with positive feeding promotion patterns documented in South Asian families [16], and the moderate Food Responsiveness (mean 2.65) and high Satiety Responsiveness (mean 3.21) CEBQ profile represents, to our knowledge, a novel baseline finding for this population: no comparable published UK South Asian infant CEBQ data could be identified in the literature for direct comparison. Future trials must deliver questionnaires digitally, in community languages, with facilitator-assisted completion, independently of PLA session delivery.

Face-to-face groups achieved higher attendance, stronger group cohesion, and higher sustainability capacity scores, with the TH Bangladeshi group recording the highest scores across most sustainability domains (mean 2.6/4). Online groups encountered technical connectivity difficulties, reduced interpersonal engagement, and incompatibility with paper-based data collection. Face-to-face delivery should be the primary mode in a definitive trial; online delivery may be retained as a contingency provided digitised data collection is embedded from the outset. The uniformly low sustainability scores in the Indian Gujarati group (mean 0.7/4) warrant further investigation and may reflect structural barriers specific to that community context.

Descriptive video analysis identified a proportional reduction in force-feeding (33% of pre-intervention videos to 8% post-intervention). However, distraction feeding remained similarly prevalent across timepoints (47% pre, 50% post), suggesting a shift in feeding strategy rather than resolution of the underlying challenge of managing reluctant eaters. This pattern warrants specific attention in future intervention content development. Interpretation is constrained by the small and unequal video samples (15 pre, 12 post) and the absence of inter-rater reliability data, which limits the rigour of the analysis. These findings are consistent with evidence from participatory and nutrition interventions for culturally and linguistically diverse populations in high-income countries, which consistently identify attendance, retention, and data collection as the primary feasibility challenges. The EIA identified no areas of adverse impact across the nine protected characteristics of the UK Equality Act 2010. Notably, 14 fathers were enrolled despite female-only eligibility criteria, reflecting the diversity of primary caregiving arrangements in this community; future trials should prospectively include all primary carers. The pilot provides an intracluster correlation coefficient (ICC) of 0.024 (95% CI −0.28 to 0.38) for BMI Z-score and a cluster-adjusted estimated sample size of approximately 159 participants per arm, informing the design of a definitive trial, notwithstanding the wide ICC confidence interval.

Strengths and limitations of this pilot warrant consideration. Key strengths include: successful enrolment across six SA ethnic and language groups in two of the most deprived boroughs in England; patient and public involvement embedded throughout design, delivery, and dissemination; structured facilitator training delivered by Women and Children First; use of validated instruments (CEBQ and PFSQ); and comprehensive item-level baseline data available for future trial planning. Methodological limitations include significant baseline imbalances across trial arms that undermine between-arm comparability; ward-level cluster randomisation insufficient to guarantee covariate balance; Waltham Forest withdrawal reducing recruitment below the planned minimum; and removal of the 12-month follow-up owing to COVID-19. Engagement limitations include session attendance of 37% and Newham retention of 48.5%, both below prespecified Go criteria; non-completion of PLA Phases 3 and 4; and a sample predominantly comprising female carers, limiting generalisability. Data quality limitations include the complete absence of follow-up questionnaire data; approximately 24% baseline questionnaire completion; no inter-rater reliability for video coding; two instrument scoring deviations identified during data analysis, affecting the CEBQ and PFSQ; and reliance on a minimum clinically important difference for BMI Z-score derived from obese-adolescent weight-management trials rather than an infant-feeding-specific threshold, since no directly applicable estimate exists for this population.

## Conclusion

This pilot demonstrated that community facilitator-led PLA interventions can be delivered within ethnically diverse SA communities in a HIC setting, with consent rates meeting prespecified feasibility thresholds and the intervention reported as universally acceptable. The pilot cannot, however, provide evidence on intervention effectiveness: the Definite Stop criterion was triggered in Newham; significant baseline imbalances preclude between-arm comparison; PLA Phases 3 and 4 were not completed by any group; and the complete absence of usable follow-up questionnaire data precludes any conclusions about the effect of NEON on child feeding behaviours or BMI Z-scores. Challenges divide into two categories requiring distinct solutions: intervention delivery challenges — session attendance of 37%, NH retention of 48.5% triggering the Stop criterion, and an incomplete PLA cycle, require reduced session burden, flexible community-integrated scheduling, and financial participant reimbursement; and outcome data challenges, i.e., approximately 24% baseline questionnaire completion, 28.4% BMI data at six-month follow-up, and failure of the eRedbook digital network diffusion platform require digitised, facilitated, multilingual data collection systems operating independently of PLA sessions and formally validated translated instruments. A definitive trial should additionally incorporate stratified randomisation by borough, ethnicity, and baseline BMI Z-score; pre-specified covariate-adjusted primary analyses; restored 12-month follow-up; and explicit progression criteria for completing all four PLA phases.

## Declarations

### Ethics Approval and Consent to Participate

Ethical approval was obtained from the UCL Research Ethics Committee (Ethics ID 17269/001; Sponsor reference: 142600; IRAS: 296259; Ethics Ref: 21/SW/0142). Participant information sheets were provided in English and community languages. Written or audio-recorded consent was obtained at least 24 hours after provision of information.

### Data Availability Statement

The data underlying the results presented in this study contain potentially identifying and sensitive information about a vulnerable participant population (mothers, fathers and infants from South Asian communities in East London) and are subject to restrictions imposed by the UCL Research Ethics Committee and the terms of participant consent. Data are not publicly available but may be requested, subject to institutional and ethical approval, from UCL via the UCL Data Protection Office, in accordance with the Data Protection Act 2018 and UK GDPR.

### Authors’ Contributions

Conceptualization: ML, LM, MH, NB, CL. Methodology: ML, LM, MH, NB, CL, PP, AF. Investigation: TK, KS, JD. Writing – original draft: AF, CAM, MH, LM. Validation, Writing – review & editing: LM, ML, MH, NB, CL, OO, JG, KWM, CI, MA, PK, KS, AF, CAM. All authors read, reviewed the study data, and approved the final manuscript.

### Collaborators

In addition to the authors, members of the NEON steering team consist of Prof Atul Singhal, Dr Sonia Ahmed, Dr Mariana Wieske, Lakmini Shah, Amelie Gonguet, Dr Zenobia Sheikh, Dr Ian Warwick, Dr Jennifer Martin, Vaikuntanath Kakarla, Dr Alex Nelson, Prof Richard Watt, Hannah Spiring, Prof Mitch Blair, Prof Audrey Prost, Dr Edward Fottrell, Ashlee Teakle, Dr Rajalakshmi Lakshman, Prof Andrew Hayward, Simon Twite, Helen Page, Chanel Edwards, Corinne Clarkson, Delceta Daley, Sumaira Tayyab, Dr Helen Crawley, Paddy Evens, Shabira Satar, Maryan Naman.

Steering team members had an opportunity to critically review results and contribute to the process of finalising this paper.

## Acknowledgements

The authors would like to thank the South Asian community facilitators in the NEON Intervention and community members of the London Boroughs of Tower Hamlet and Newham for their important contribution and engagement to this research project.

We would like to express our gratitude to Shereen Al Laham for her invaluable assistance in the initial setup of the study design and ethical considerations. Furthermore, we would like to acknowledge the contribution of the NEON Core Team, Steering Team and all health experts who contributed to this study and validating the intervention and NEON toolkit. We want to thank the Women & Children First Charity and First Steps Nutrition Trust for their valuable contributions and guidance throughout the study. Steering team members had an opportunity to critically review results and contribute to the process of finalising this paper. The authors would like to thank the National Institute of Health Research (NIHR) Academy and the NIHR Collaboration for Leadership in Applied Health Research and Care North Thames for funding the NEON study. This work is also supported by the NIHR GOSH BRC. The views expressed are those of the author(s) and not necessarily those of the NHS, the NIHR or the Department of Health.

## Participant consent for publication

Participant information sheets and consent forms were provided to community members and those expressing interest. Community participants agreed to participate and gave audio/video consent prior to their participation in the workshops. Verbal consent was witnessed and formally recorded. All participants were informed of their right to freely withdraw from the study at any time. Confidentiality of personal data was ensured through the use of anonymisation techniques as stated in the Data Protection Act (1998) and in line with the General Data Protection Regulation (2018). All participant data is anonymised and stored on an encrypted password protected computer. Data can only be accessed by the authorised research personnel.

## Trial registration

ISRCTN10234623 (IRAS ID: 296259; Ethics Ref: 21/SW/0142). Prospectively registered.

## Supporting information

**S1 Protocol.** NEON pilot cluster randomised controlled trial study protocol. Previously published as: Manikam L, Allaham S, Patil P, Naman M, Ong ZL, Demel IC, et al. NEON PLA women’s groups to improve infant feeding in South Asian infants: pilot RCT study protocol. BMJ Open.

2023;13:e063885 [16]. (PDF)

**S2 Checklist.** CONSORT 2010 checklist, with the 2016 extension to randomised pilot and feasibility trials. (DOCX)

